# Phylogenetic analysis of streptococci in samples taken from the throat cultures of children in Turkey and the presence of mef(A), mef(E), erm(B) and erm(TR) genes in patients with *Streptococcus pyogenes*

**DOI:** 10.1101/2020.09.22.20196410

**Authors:** Çiğdem Eda Balkan, Hayrunisa Bekis Bozkurt, Cem Öziç

## Abstract

**Objective:** Failure to achieve success with penicillin treatment in some cases observed in the pediatric group and the decrease in macrolide activity have brought about the necessity of a new study aiming to differentiate bacteria at the species level in throat infections. Antibiotic resistance studies are of great importance for the treatment of bacterial infections in terms of public health and rational antibiotic use. For this purpose, we aimed to perform a species-level differentiation of streptococci isolated from the throat cultures of pediatric patients presenting to our hospital, to determine their antibiotic susceptibility, and to identify the macrolide resistance genes of mef(A), mef(E), erm(B) and erm(TR) in patients with *Streptococcus pyogenes*. The study included 51 samples taken from pediatric cases presenting with a sore throat as the patient group and 36 samples from children without this complaint as the control group.

**Material and Method:** The throat culture samples taken from 51 children presenting to the hospital with the complaint of sore throat were evaluated in the laboratory, and streptococcus was diagnosed using tests; gram staining, catalase and PYR, and the susceptibility profile was determined with the Kirby-Bauer disk-diffusion method. Bacteria were identified at the species level according to 16srRNA sequences, and possible macrolide resistance genes of mef(A), mef(E), erm(B) and erm(TR) were determined by PCR in species detected to have *S. pyogenes*.

**Results:** Our antibiotic susceptibility results were consistent with the general results reported in Turkey. The sequence analysis of bacteria was performed according to 16srRNA sequences, and *S. pyogenes, Streptococcus pneumoniae, Streptococcus anginosus, Streptococcus agalactiae*, and *Streptococcus dysgalactiae* were isolated. In patients with *S. pyogenes*, the genetic determinants of macrolide resistance, mef(A), mef(E), erm(B) and erm(TR), were investigated with the PCR method using primers specific to each gene. Different levels of expression were observed in five patients. Macrolide resistance in *S. pyogenes*, which is reported at various percentages in the world, was found to be 9.8% in our study.

**Discussion:** The results of our study show that penicillin resistance genes were found in five of the patients evaluated. When the anamnesis of these patients was examined, it was determined that there were patients that frequently presented to the hospital with throat infections and experienced re-infection within a few weeks after receiving treatment. The common discourse of clinicians is that there may be an unknown resistance development. Therefore, our research should be supported by new hypotheses and studies that are open to development.

## Introduction

*Streptococcus pyogenes* beta hemolytic is a gram-positive bacterium included in Group A streptococcus (GAS) according to the Lancefield classification and has caused infections for centuries. Although it is associated with a wide range of diseases that can progress from skin infections to sepsis, it is largely known as being the most common infectious agent in throat and upper respiratory tract infections. [1] In addition to causing acute diseases, it can also lead to serious complications, such as acute rheumatic fever and glomerulonephritis. Especially in newborns, it is observed that the frequency of infection increases after the gradual decrease of immunity acquired through breast milk. In schoolchildren (aged 5-15 years), the incidence of GAS pharyngitis is not negligible, with the presence of GAS being asymptomatic in about 15-20% of this age group. [2] It is very important to detect bacteria to eradicate the disease and prevent associated complications with antibiotic treatment, as well as stopping its spread, especially in environments with a high contagion risk, such as schools and nurseries. [3-4]

With the discovery of penicillin for which Fleming, Florey and Chain received the Nobel Prize in 1945, the world has gained great power in the fight against bacteria. The use of penicillin as the first-choice treatment for GAS infections and other gram-positive bacteria continues, except for certain species that have become resistant by natural selection. [4,5] For many years, macrolides have been used as a second option when there is no response from penicillin treatment against gram-positive bacteria. Despite this situation being commonly observed in practice, there is no study conclusively reporting that streptococci have developed resistance to penicillin, except for a few limited publications showing the reduced susceptibility of these bacteria. In some clinical studies, it has been observed that 30% of the patients did not respond to penicillin treatment in tonsillopharyngitis caused by GAS. This situation is considered to occur as a result of the evolution of bacteria, especially in terms of their mechanisms of escape from antibiotics. [6] Macrolides, which are used as the second treatment choice in patients, are also globally becoming increasingly resistant to antibiotics. [7] For this reason, it is of great importance to detect virulence factors, antibiotic escape mechanisms, microorganism subtypes, and resistant genes of GAS infections. As is known, M protein is among the main virulence factors in GAS, and strains detected with the use of M antisera in species differentiation are called ‘M serotypes’. However, today, it is known that there are streptococcal groups that cannot be defined based on M protein alone. For this reason, genotype determination and especially 16s rRNA are used in the differentiation of streptococci and other bacteria. In this process, the term ‘emm type’ is used to refer to strains identified by sequencing. [8] According to the conducted studies, there is often unnecessary antibiotic use without a culture analysis in infections for which GAS bacteria are possibly responsible. [9] Despite seeming simple, it is actually very difficult to eradicate GAS infections considering their clinical implications while also avoiding unnecessary antibiotherapy and preventing resistance to antibiotics. [9] To date, there is no vaccine for this bacterium, and this presents a serious risk in certain conditions of neonatal sepsis and infant mortality. [10]

In our study, throat culture samples taken from 51 children that presented to our hospital with the complaint of sore throat were evaluated in the laboratory, a diagnosis of streptococcus was made, and their susceptibility profiles were obtained. It was aimed to perform a differentiation at the species level according to 16srRNA sequences and investigate the possible presence of resistance genes in species identified as *S. pyogenes*.

## MATERIAL AND METHOD

### Kirby-Bauer Disk-Diffusion Method

Of the 78 throat culture samples taken from the pediatric patients presenting to our hospital with throat infections in our region, 27 of those belonging to the same patients were excluded, and the remaining 51 were placed in the Cary Blair transport medium using swabs and transferred to the microbiology laboratory. The samples were first inoculated onto blood agar and then kept in an oven for 24 hours, after which the PYR test was applied to beta hemolytic colonies. Using the Kirby-Bauer disk-diffusion method, beta hemolytic bacteria adjusted to 0.5 MacFarland standard were seeded on blood agar, and Cefoxitin, Ciprofloxacin, Levofloxacin, Gentamicin, Amikacin, Netilmicin, Teicoplanin, Vancomycin, Erythromycin, Clindamycin, Tetracycline, Linezolid, Rifampin, Trimethoprim-Sulfamethoxazole, Cefdinir, Cefixime, Cefotaxime and Bacitracin disks were placed on the medium. The susceptibility limits of antibiotics according to the Eucast criteria are given in Table 1 The swabs transported in the Carry Blair transport medium were stored at -80 °C. [11]

**Table 1.**
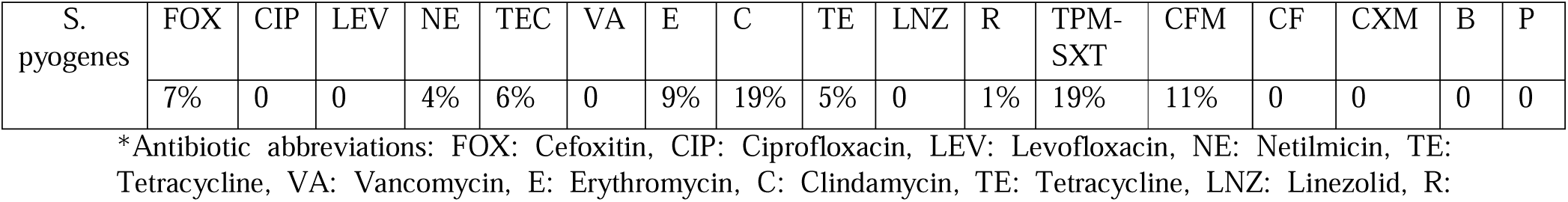
Percentage of antibiotic resistance of patients with *S. pyogenes* using the Kirby-Bauer disk-diffusion method according to the EUCAST criteria

### Genomic DNA Isolation Protocol

The samples were placed into Eppendorf tubes, to which 200μl dH2O, 50μl 0,5M EDTA, 10μl %20 sarkosyl, 10μl proteinase K (10mg/ml), 10μl 1M Tris-HCl (pH:8), and 5μl 5M NaCl were added. The mixture was vortexed for 5 min and kept in a water bath set at 65 °C for 30 min. During this period, the mixture was vortexed every 10 minutes. Phenol: chloroform: isoamyl alcohol (25:24:1) was added to the cell suspension. It was then centrifuged at 13,000 rpm for 5 min. The supernatant layer was removed with a Pasteur pipette (or a 1,000 μl micropipette the tip of which was cut with a razor blade) and transferred to a new tube. The phenol: chloroform: isoamyl alcohol procedure was performed three times as described above. In each step, the supernatant was removed from the products obtained at the end of centrifugation and transferred to a clean Eppendorf tube, to which 3M NaAc at 1/10 of its volume and absolute ethanol at two times of its volume were added, and this mixture was kept overnight at -20 °C. At the end of this period, the sample was centrifuged at 13,000 rpm for 10 min. The supernatant was removed, and the pellet was dried. After adding 200μl dH2O, the pellet was thawed and 0.3M NaOAc and 440μl ethanol were added at 1/10 volume and kept overnight at -20 °C. At the end of this period, the sample was centrifuged at 13,000 rpm for 5 min. The supernatant was removed, and the pellet was allowed to dry. After drying, the pellet was thawed in 100 µl dH2O. Genomic DNA obtained was examined for quality, RNA contamination and integrity according to the spectrophotometric measurement first, followed by imaging in 0.8% agarose gel. [12]

### Gene Sequences used for the Detection of Resistance Genes

In this study, primers containing the following gene sequences were used. [13]

erm(B): f 5’-ATTGGAACAGGTAAAGGGC-3’ and r 5’-GAACATCTGTGGTATGGCG-3’

erm(TR): f5’-ACAGAAAAACCCGAAAAATACG-3’ and r5’ TTGGATAATTTATCAAGATCAG-3’

mef(A): f 5’-TGGTTCGGTGCTTACTATTGT-3’ and r 5’-CCCCTATCAACATTCCAGA-3’

mef(E): f 5’-GGGAGATGAAAAGAAGGAGT-3’ and r 5’-TAAAATGGCACCGAAAG-3’.

### Genomic DNA Replication

Genomic DNA was used as source DNA in the PCR reaction. The PCR reaction was established with 16S rRNA (F and R) primers. A solution containing 2.5 µl 10X buffer, 2.5 µl 25 mM MgCl2, 2 µl 2.5 µM dNTP mixtures, 2.5 µl F, 2.5 µl R, 0.5 µl genomic DNA, and 0.2 µl Taq DNA polymerase enzyme (5u/µl) was completed to a total volume of 25 µl by adding 12.3µl ddH2O. The PCR program used for products was as follows: at 94 °C for 2 min, at 94 °C for 1 min, at 55 °C for 1 min, at 72 °C for 1 min, at 72 °C for 4 min, and at 4 °C for ∞.

### Agarose Gel Analysis And Gel Imaging

The DNA fragments run in the agarose gel were checked in a UVP transilluminator device, and the data were recorded using a UV-photometer gel documentation device (UviTec).

### Purification of PCR Products From Gel

The sizes of the banding formed in the agarose gel in the products as a result of PCR were determined using appropriate markers. The bands suitable for the expected size were removed from the gel. Then, purification was carried out using the QIAquick gel extraction kit and protocol according to the user manual of the kit. The sequence analysis process was performed as required for the determination of purified samples species. The results were evaluated with the BLAST analysis using the NCBI database, and the similarities between the species were determined. A phylogenetic tree was created using CLUSTALW2 and Mega5.

## RESULTS

### Antibiotic Susceptibility Results

Antibiotic susceptibility was identified in 51 throat swabs taken from pediatric patients presenting to our hospital with throat infections (Table 1).

The antibiotic resistance rates of all streptococcus species were determined as follows: 37% for Cefoxitin, 11% for Netilmicin, 16.2% for Teicoplanin, 24.3% for Erythromycin, 25% for Clindamycin, 13.5% for Tetracycline, 2.7% for Rifampicin, 24.2% for TPM-SXT, 12% for Cefdinir, 22.7% for Ampicillin, and 17% for Penicillin. There was no resistance to Cefixime, Vancomycin, Linezolid, Ciprofloxacin, and Cefotaxime.

### Acquisition of Genomic DNA

The genomic DNAs of all bacteria identified in the study were isolated. The DNA concentrations were determined spectrophotometrically, and the working concentration was adjusted. The DNAs obtained as a result of genomic DNA isolation were amplified with primers specific to the 16S rRNA gene regions, and the products formed as a result of PCR were imaged with gel electrophoresis. These products were run in gel electrophoresis, and it was observed that 1300 base pairs were formed, as expected for size. The 16S gene was obtained by PCR. The PCR results of the patient and control groups are shown in Figures 1 and 2, respectively.

**Figure 1.**
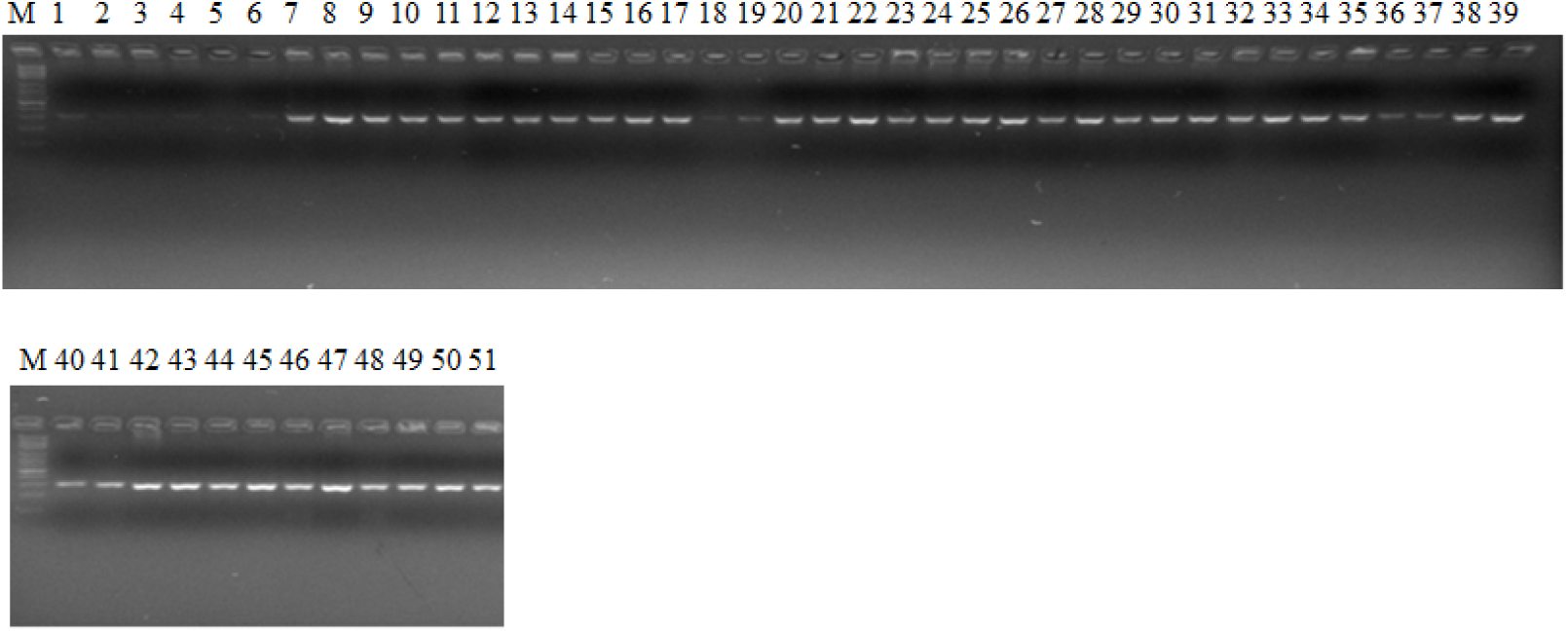
PCR image of the patient group.

**Figure 2.**
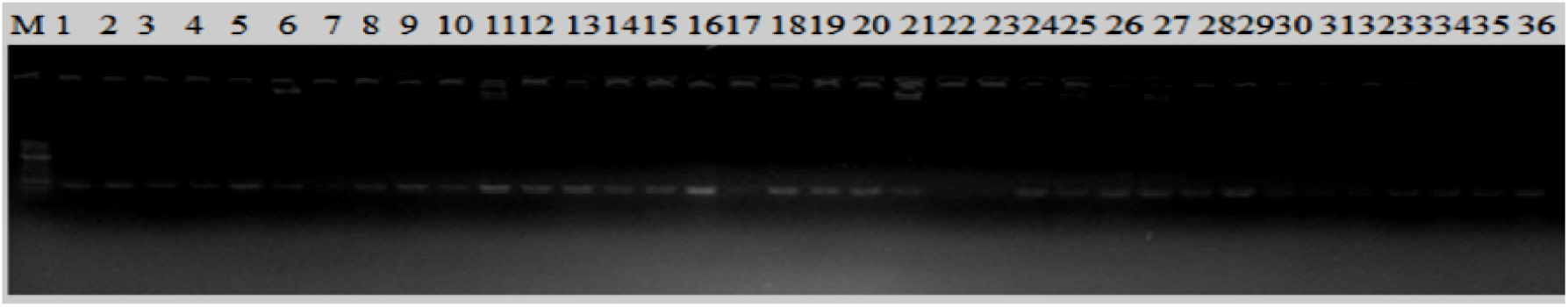
PCR image of the control group.

The sequence analysis was performed on the products obtained by PCR, and as a result, five different *Streptococcus* species were identified. In Tables 2 and 3, the species identified according to the 16s rRNA analysis are coded with numbers to maintain patient confidentiality.

**Table 2.**
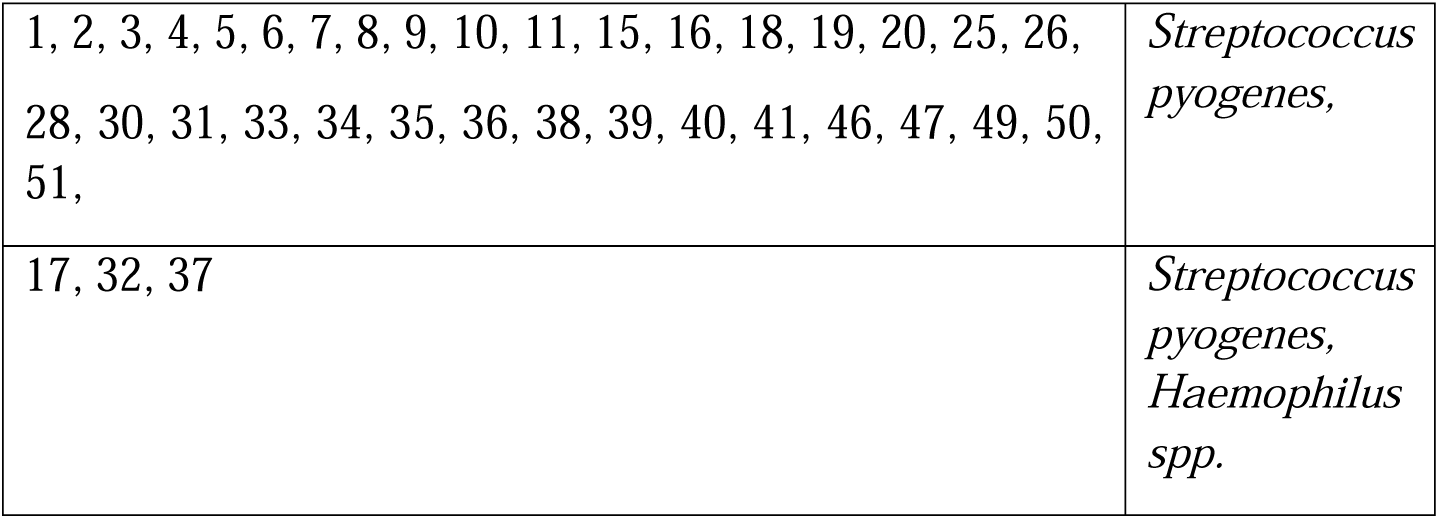
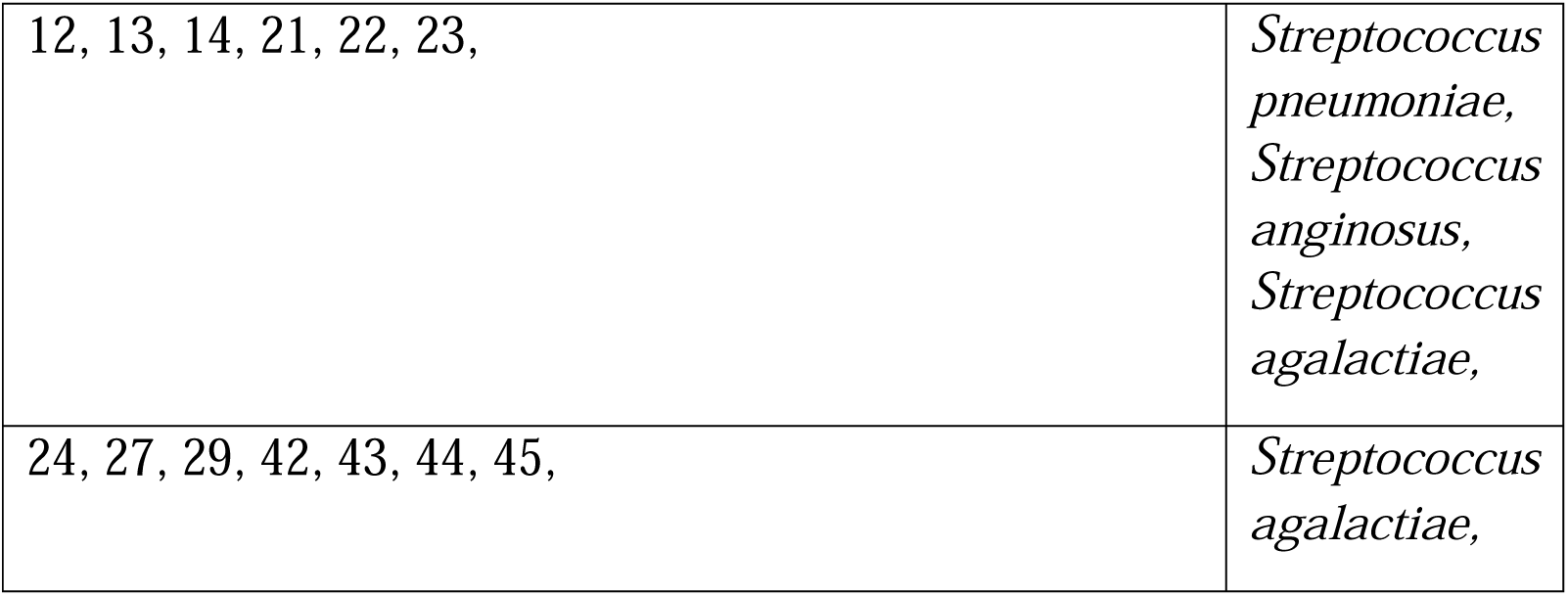
16S rRNA results of the patient group

**Table 3.**
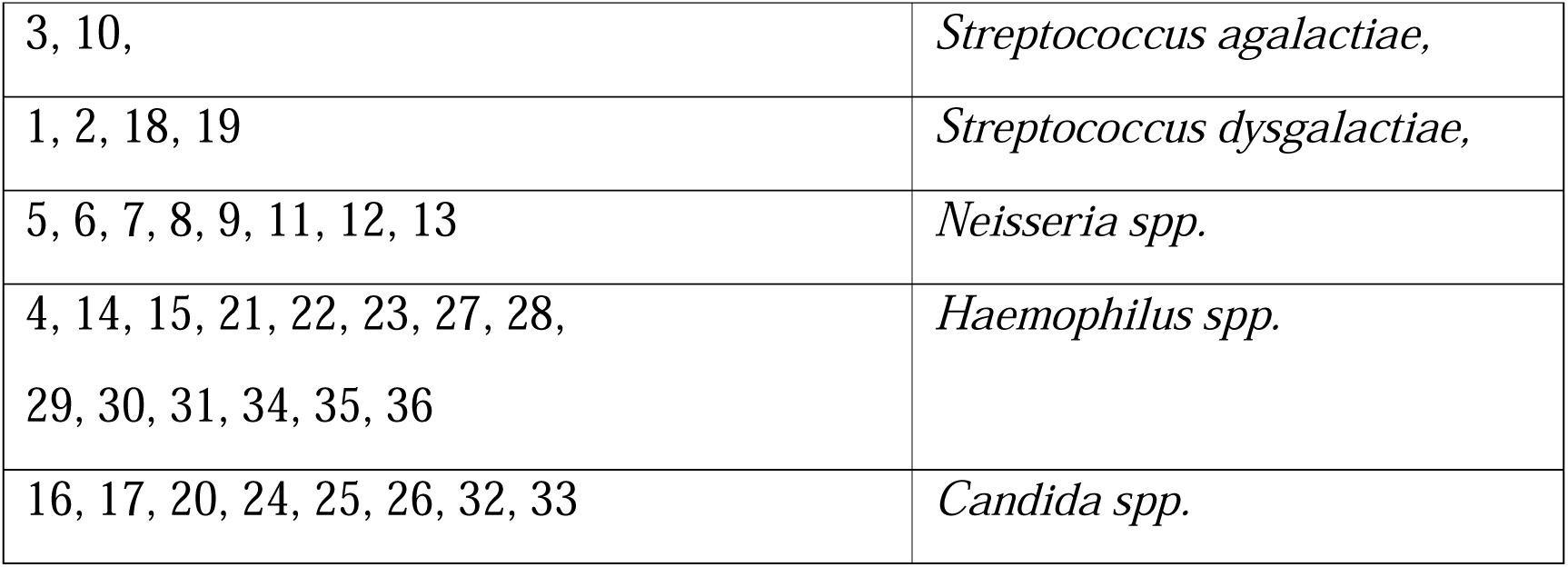
16S rRNA results of the control group

The species generally identified within the scope of the study were *S. pyogenes, Streptococcus pneumoniae, Streptococcus anginosus, Streptococcus agalactiae*, and *Streptococcus dysgalactiae*. A phylogenetic analysis was performed using the results of the sequence analysis. Figure 3 presents the species identified according to the phylogenetic analysis.

**Figure 3.**
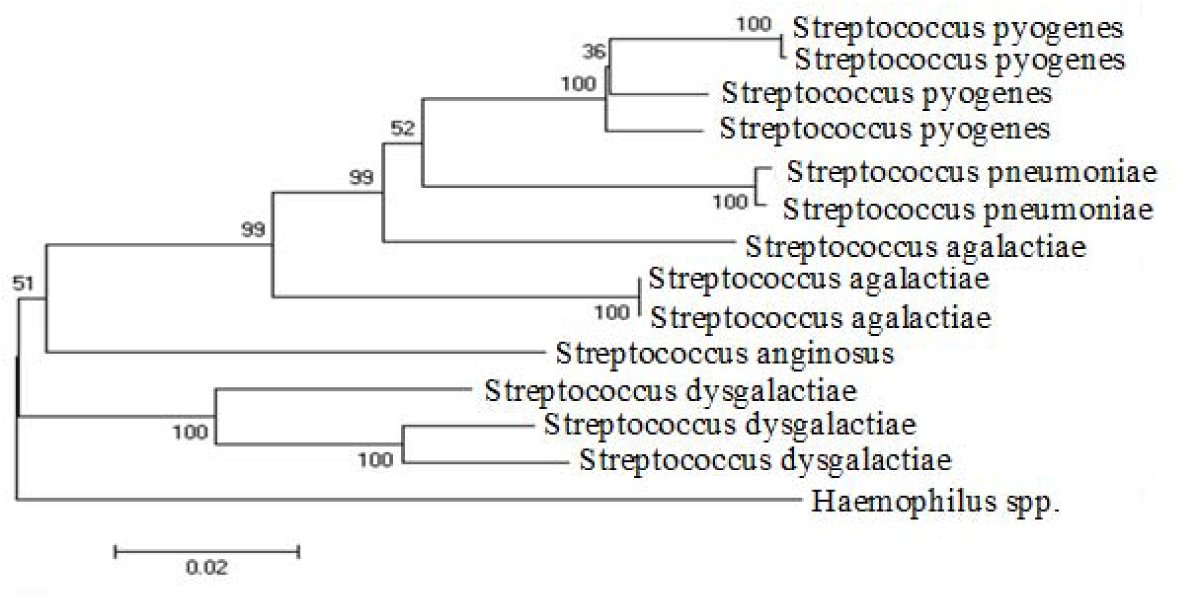
Results of the phylogenetic analysis of the species.

### Resistant Gene Regions

In this study, we aimed to determine the penicillin resistance of the identified species. The genetic determinants of macrolide resistance were investigated with the PCR method using primers specific to each of the mef(A), mef(E), erm(B) and erm(TR) genes. Within the scope of the study, it was observed that the expression levels of these genes decreased in some patients (n = 5), and this was considered to be the reason for the lack of response to penicillin and macrolides in these cases (Figure 4).

**Figure 4.**
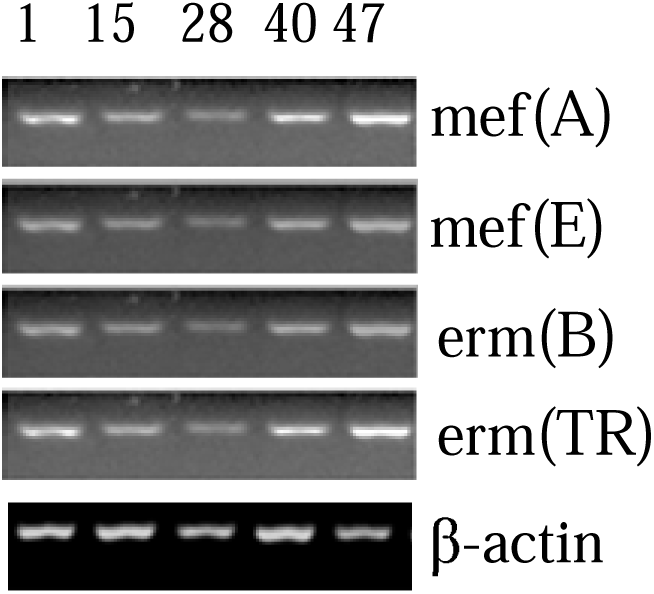
Agarose gel image of the PCR results of the mef(A), mef(E), erm(B) and erm(TR) genes in five patients (Patients 1, 15, 28, 40 and 47) showing resistance.

### Discussion

GAS is the most common cause of throat infections among bacteria. Both acute and chronic forms of GAS can be seen at the age of three and over; therefore, they are diseases that frequently occur in a wide age group. [14] Contagion occurs with close contact through infected droplets. In addition to domestic transmission, other environments with a high risk of transmission include public areas, such as schools, barracks, and kindergartens. [15] Especially in acute forms, the disease usually spontaneously regresses in a week without the addition of antibiotics to treatment. However, in the presence of severe complications, including acute rheumatic fever and glomerulonephritis, it is recommended to start penicillin treatment without waiting for the culture result, and Hanage et al. stated that resistant strains were developing in patients that did not respond to treatment. [16] Since studies began to be conducted in 1985, growing experience in clinical practice indicates that some patients have resistance against penicillin treatment and that it does not seem beneficial in eradicating Group B streptococci, but this situation cannot be proven in vitro in real life. [17] Examining the results of some antibiotic resistance studies across the world, Erythromycin-resistant *S. pyogenes* was found in 35% of the children admitted to hospital with throat infections in Italy. [18] Similarly, in France, Erythromycin resistance was reported at a rate of 6.2% in the throat cultures of 1,500 pediatric patients. [19] In our study, the rate of Erythromycin resistance was determined as 9%, which is similar to previous studies. It is also seen that there is gradually increasing macrolide resistance inversely proportional to age, especially among pediatric patients. In a study conducted in Germany, a Group B streptococcus strain that was not susceptible to penicillin was isolated from a single patient. [20] A similar report has not yet been published in Turkey. Examining the in vitro effects of antibiotics, we obtained similar results to the literature concerning the distribution and antibiotic susceptibility of *S. pyogenes* strains in the pediatric group. [21,22] However, in contrast to previous studies, we also performed a differentiation of strains at the species level in a patient group from our region presenting to our hospital with a sore throat, investigated their antibiotic susceptibility, and determined whether there was any resistance by examining the mef(A), mef(E), erm(B) and erm(TR) genes among the cases that did not respond to penicillin treatment and were suspected to have macrolide resistance.

When the distribution of resistance among our patients was examined, it was observed that resistance to Clindamycin in *S. pyogenes* was 19% while it reached a higher level (25%) considering all streptococcal species. In a study undertaken by Oryaşin et al., it was reported that the erm B and erm TR genes might have this activity of resistance. [23] In the control group in our study, we observed normal throat flora elements, such as *S. agalactiae, S. dysgalactiae, Neisseria* spp. *Haemophilus* spp. and *Candida* spp.

Globally, there are many studies showing the development of penicillin resistance to Group B streptococci (*S. agalactiae*). The first *S. agalactia* resistant to penicillin was reported in 1987, followed by similar reports in many other countries, and the aacA-aphD gene was held responsible for this resistance. [24] To prevent maternal transmission from mother to baby, it is recommended that antibiotic prophylaxis should be applied before normal delivery. [25] Although clear penicillin resistance has not yet been reported for *S. pyogenes*, macrolides are used as a second option in patients who do not respond to penicillin treatment. [26] In an animal study by Samir et al., penicillin- and macrolide-resistant *S. pyogenes* was identified, and ermB, one of the resistance genes, was detected in the whole sample. The authors commented that this situation posed a risk of bacterial transmission to humans through children that are in close contact with animals. [27] In another study, Vannice et al. showed the effect of pbp2x point mutation reducing sensitivity to penicillin in a case series of two patients. [28] In our study, resistance genes were found at various expression levels in five patients in a sample of 51 patients. According to the hospital records, our constituted a population that presented to the hospital several times a year due to throat infections. It was observed that the patients in this group did not respond well to macrolide treatment. It is considered that *S. pyogenes* strains, which have not yet received as much global attention as they require, may gradually become more resistant, to the extent of being described as super-resistant in future. [16]

Some studies have also mentioned the necessity to use secondary treatment options in throat infections that do not respond to penicillin treatment and emphasized how wrong it was to prescribe medicine for pediatric and adult patients by considering their symptoms alone. [29] In the vast majority of studies, macrolide resistance genes were found at various levels. As stated in the literature on this subject, there are various differences between countries even in relation to the structures of resistance genes. [30] In a study conducted in Norway, the mef A gene found in *S. pyogenes* was observed to differ from that found in *S. pneumonia*, and this gene was noted to have many subtypes. [31] In another Norwegian study, it was stated that the erm TR gene was present in 26 of 44 Erythromycin-resistant strains, erm B or erm TR in six, and mef E in one. [32] Resistance genes, mechanisms and increased resistance in *S. pyogenes*, as in all bacteria, cause great economic and moral losses across the world. According to the European Centre for Disease Prevention and Control reports, millions of liras are spent every year for this infection, which easily spreads among children and often requires antibiotic treatment. [33] Nevertheless, there are yet-to-be-proven efficacy problems concerning the first treatment option, penicillin, while at the same time, macrolides, one of the primary alternatives, is also becoming more resistant with each passing day. [34]

## CONCLUSION

The results of our study showed the presence of various resistance genes in five of the patients evaluated. When the anamnesis of these patients was examined, they were seen to represent a pediatric group that visited the hospital due to frequent, long-lasting throat infections and experienced re-infection within a few weeks after receiving treatment. Decreased penicillin susceptibility in *S. pyogenes* is a concept that has not yet been proven due to the availability of only a limited number of studies. Similarly, due to their reduced susceptibility, macrolides can occasionally be inadequate in eradicating this infection, which seems simple but incurs serious health-related cost across the world. Further comprehensive studies are required to initiate radical changes in the approach of countries to throat infections. Our study should be supported by new antibiotics resistance studies designed for this purpose and open to development.

## Data Availability

All data referred to in the manuscript available.

## Acknowledgment

This study was financially supported by the Scientific Research Projects Unit of XX University, with the project number 2018-TS–67.

## Notes

### Competing Interest Statement

The authors have declared no competing interest.

### Funding Statement

This study was financially supported by the Scientific Research Projects Unit of Kafkas University with the project number 2018TS67

### Author Declarations

8057635-050-99/88 Kafkas University Medical Commitee Ethical approval number.

